# Effectiveness of an eHealth Intervention for Reducing Psychological Distress and Increasing COVID-19 Knowledge and Protective Behaviors among Ethnoracially Diverse Sexual and Gender Minority Adults: A Quasi-experimental Study (#SafeHandsSafeHearts)

**DOI:** 10.1101/2023.01.09.23284380

**Authors:** Peter A. Newman, Venkatesan Chakrapani, Notisha Massaquoi, Charmaine C. Williams, Wangari Tharao, Suchon Tepjan, Surachet Roungprakhon, Joelleann Forbes, Sarah Sebastian, Pakorn Akkakanjanasupar, Muna Aden

## Abstract

**Purpose:** Lesbian, gay, bisexual, transgender, queer, and other persons outside of heteronormative and cisgender identities (LGBTQ+) and ethnic/racial minority populations are at heightened vulnerability amid the Covid-19 pandemic. Systemic marginalization and resulting adverse social determinants of health contribute to health disparities among these populations that result in more severe consequences due to Covid-19 and the public health measures to control it. We developed and tested a tailored online intervention (#SafeHandsSafeHearts) to support ethnoracially diverse LGBTQ+ individuals in Toronto, Canada amid the pandemic.

**Methods:** We used a quasi-experimental pre-test post-test design to evaluate the effectiveness of a 3-session, peer-delivered eHealth intervention in reducing psychological distress and increasing Covid-19 knowledge and protective behaviors. Individuals ≥18-years-old, resident in Toronto, and self-identified as sexual or gender minority were recruited online. Depressive and anxiety symptoms, Covid-19 knowledge and protective behaviors were assessed at baseline, 2-weeks postintervention, and 2-months follow-up. We used generalized estimating equations and zero-truncated Poisson models to evaluate the effectiveness of the intervention on the four primary outcomes.

**Results:** From March to November 2021, 202 participants (median age, 27 years [Interquartile rage: 23-32]) were enrolled in #SafeHandsSafeHearts. Over half (54%, n=110) identified as cisgender lesbian or bisexual women or women who have sex with women, 26.2% (n=53) cisgender gay or bisexual men or men who have sex with men, and 19.3% (n=39) transgender or nonbinary individuals. The majority (75.7%, n=143) were Black and other people of color. The intervention led to statistically significant reductions in the prevalence of clinically significant depressive and anxiety symptoms, and increases in Covid-19 protective behaviors from baseline to postintervention.

**Conclusion:** We demonstrated the effectiveness of a brief, peer-delivered eHealth intervention for ethnoracially diverse LGBTQ+ communities in reducing psychological distress and increasing protective behaviors amid the Covid-19 pandemic. Implementation through community-based health services with trained peer educators supports feasibility, acceptability, and the importance of engaging ethnoracially diverse LGBTQ+ communities in pandemic response preparedness. This trial is registered with ClinicalTrials.gov, number NCT04870723.

## Introduction

Marginalization of lesbian, gay, bisexual, transgender, queer (LGBTQ) and other persons outside of heteronormative and cisgender identities (LGBTQ+), and ethnoracially diverse populations, exacerbates vulnerability to SARS-CoV2 transmission and serious Covid-19 outcomes. Adverse social determinants of health (SDOH) owing to systemic discrimination against LGBTQ+ [1-4], Black [5] and other racial/ethnic minority populations [6,7], and those with intersectional marginalized identities [2,3,8], contribute to health disparities and excess risk in the Covid-19 pandemic. Among LGBTQ+ people in Canada, precarious employment and financial insecurity [9,10], unstable housing [10], lack of LGBTQ+-competent healthcare [11]—even more so among racial and ethnic minorities, and women [12]—and pervasive stigma [1] increase risks for Covid-19 transmission, reduce access to care, and constrain the ability to adhere to public health-recommended nonpharmaceutical interventions, such as physical distancing, working from home, and Covid-19 testing.

Health and mental health disparities among LGBTQ+ and ethnoracially diverse people associated with social and structural vulnerabilities are likely to be exacerbated by the trauma and social isolation of the Covid-19 pandemic [1,3,13-16]. Evidence from Canada [17-19] as well as the U.S. [8,20] indicates not only increases in, but disproportionately higher rates of depression, anxiety, and loneliness among LGBTQ+ people compared to cisgender heterosexual individuals during the pandemic. Yet, despite documented health disparities and heightened vulnerability among LGBTQ+ and ethnoracially diverse populations, we are aware of no tailored, evidence-informed interventions to reduce the burden of Covid-19 among these communities.

We developed and tested #SafeHandsSafeHearts, a peer-delivered eHealth intervention, with the aim of providing psychosocial and behavioral support and education to ethnoracially diverse LGBTQ+ individuals during the Covid-19 pandemic. We also aimed to advance pandemic responses designed for ethnoracially diverse LGBTQ+ populations.

## Materials and Methods

This study was funded by the International Development Research Centre (109555) and the Social Sciences and Humanities Research Council of Canada (895-2019-1020). All study procedures were approved by the Research Ethics Board of the University of Toronto (protocol no. 39769) and conducted in accordance with the Declaration of Helsinki. Informed consent was obtained from all participants. The clinical trial was registered on ClinicalTrials.gov (NCT04870723). Recruitment of participants for baseline, postintervention, and follow-up assessments lasted from March 1, 2021 to May 15, 2022. The study was registered prospectively as one site of an international randomized clinical trial with a crossover design; however, due to protracted delays in ethics approvals from the other two sites amid a public health emergency, we implemented the approved study at the Toronto site as a pilot using a quasi-experimental pretest-posttest design. All enrollment criteria remained the same, and the intervention and assessments were conducted according to the registered protocol. The authors confirm that all ongoing and related trials for this intervention are registered. The supporting CONSORT checklist is available as supporting information; see S1 Checklist.

### Study Design and Setting

We used a quasi-experimental, single group pretest-posttest design to assess the effectiveness of the #SafeHandsSafeHearts eHealth intervention. The study was wholly developed and conducted online due to rolling lockdowns in Toronto and the contiguous urban region from March 2020 to June 2021 [21]. The Greater Toronto and Hamilton Area (GTHA; pop. estimate, 7.3 million) is comprised of the largest and ethnoracially diverse cities by population in Ontario, including Toronto, Halton, Peel, York, and Durham. We used a World Health Organization-recommended approach [22] with community engagement in intervention development, co-governance, and capacity-building of community-based organizations (CBOs). Given the disproportionate impact of the Covid-19 pandemic on physical and mental health among LGBTQ+ and ethnoracial minority populations [3,13,14,20], and as a community-based intervention, we used a single group design rather than randomization to a control or waitlist group during a public health emergency.

### Sample Size

The sample size was calculated based on power (80%) to detect significant differences (alpha = 0.05 for 95% confidence interval, two-tailed test) in 4 primary outcomes: increases in Covid-19 knowledge and Covid-19-protective behavior scores; and decreases in the proportion of participants with pandemic-related depressive and anxiety symptoms. We assumed a small to medium effect size (Cohen’s d = 0.3) on knowledge and behavior, and 20% reduction in the proportion of participants with depression or anxiety. Required sample sizes estimated using Stata-16 and G*Power 3.1 ranged from 90–92 to detect significant differences between pre-intervention and postintervention timepoints. Assuming 10% attrition and adding a design/clustering effect of 1.5, the target sample size was increased to 239.

### Procedures

We developed a customized online dashboard AND database to support study coordination, including tracking of participant recruitment, screening, enrollment, counselor assignment, and eHealth sessions. Participants were recruited online via listservs and social media accounts of CBOs and health centers serving ethnoracially diverse LGBTQ+ communities, LGBTQ+ e-groups, and a study website. We distributed e-flyers and messages with a focus on groups and organizations serving ethnoracial minority LGBTQ+ populations.

Eligibility criteria were (1) age ≥18 years; (2) self-identify as cisgender lesbian or bisexual woman or woman who has sex with women (LBWSW); cisgender gay or bisexual man or man who have sex with men (GBMSM); or transgender or non-binary individuals (TNB); (3) resident in the GTHA for ≥ 6 months; (4) able to understand and willing to provide informed consent; and (5) able to understand English.

Peer counselors were recruited among the focal study populations and those with experience working with LGBTQ+ and ethnoracially diverse communities from social service organizations and social work training programs. Peer counselors received a 3-day, manualized online training, including small-group discussions, role-plays, mock counseling sessions, and feedback, delivered by study coordinators and health and mental health professionals, as well as a subsequent booster session. The training covered Covid-19, public health-recommended nonpharmaceutical interventions, pandemic-related distress (i.e., anxiety, depression, social isolation), motivational interviewing (MI)-based counseling, psychoeducation, and research ethics. Biweekly clinical group supervision was conducted throughout the intervention, along with two Covid-19 updates provided by Toronto Public Health medical staff.

### Intervention

#SafeHandsSafeHearts builds on evidence-informed interventions that have used MI [23,24] and psychoeducation [25] to increase health knowledge, health behaviors, and reduce psychological distress [26-28]. Both MI and psychoeducation lend themselves to culturally appropriate, nonjudgmental, non-stigmatizing and strengths-based approaches to self-evaluation, emotional support, and behavior change [29,30], and have demonstrated effectiveness with LGBTQ+ and racial/ethnic minority populations [30-32].

The intervention content was organized into 3 sessions (see S2 Appendix). The first session focused on building rapport, goal-identification (i.e., participant’s goals for change), and psychoeducation (i.e., selecting 1 or 2 new things to observe/try out and discuss next session), with additional content focused on knowledge about Covid-19 transmission, symptoms, testing, and treatment. Session 2 content focused on understanding and practicing Covid-19 protective behaviors (i.e., improving self-efficacy), risk reduction, psychoeducation, and problem-solving (i.e., reviewing the “new things” experience, reinforcing successes, normalizing setbacks). Session 3 focused on understanding psychosocial issues, promoting awareness of community resources, and improving mental health (i.e., mental health and social support assessment and strengthening, expanding support for change—tools, relationships, services), and maintaining change (i.e., relapse prevention).

The intervention was delivered online (via mobile phone, tablet, laptop, or PC) in 3-weekly 60-minute modules. Counselors were given flexibility to address pressing needs raised by participants at the beginning of each session. A referral list was provided to all counselors, including locally available and free or low-cost concrete services (e.g., food banks, housing), health services, and mental health hotlines, including LGBTQ+- and ethnoracially-competent organizations. Counselors documented referrals made in each session and followed up with participants in subsequent sessions. Participants were provided with a $30 honorarium after each online session, and after postintervention and follow-up survey completion.

### Measures

Participants completed surveys at baseline, 2-weeks postintervention after their final eHealth session, and 2-month follow-up.

#### Primary outcomes

Depression symptoms in the past 2 weeks were measured using the Patient Health Questionnaire-2 (PHQ-2) [33]. Anxiety symptoms in the past 2 weeks were measured using the Generalized Anxiety Disorder-2 scale (GAD-2) [34]. Each 2-item scale was scored from 0–3 (range, 0–6), with a score of ≥3 indicative of screening cut-points for clinically significant depression or anxiety [35].

Covid-19 knowledge was assessed using a 7-item index (score range, 2–7) developed by the research team, based on U.S. Centers for Disease Control (CDC) guidelines [36,37] and published research [38,39]. Public health-recommended Covid-19 protective behaviors (handwashing, mask-wearing, physical distancing) were assessed using a 9-item index (score range, 1–18) developed by the research team based on WHO and U.S. CDC guidelines [36,37].

#### Covariates

Demographic variables included age, sex, gender identity, sexual orientation, race/ethnicity, city of residence, country of birth, education, and employment status. We assessed loneliness/social isolation (UCLA Loneliness Scale) [40]; Covid-19 stress (Covid-19 danger and Covid-19 traumatic stress subscales) [41]; resilience (Resilience to Traumatic Experience scale) [42]; vaccine conspiracy beliefs [43-45]; and Covid-19 vaccination status.

### Acceptability of the Intervention

After each eHealth session, participants were sent an online link to a confidential 4-item evaluation to indicate their satisfaction with the session and session length, the extent to which it was helpful in improving their emotional wellness or mental health and their Covid-19-related knowledge and skills.

### Statistical Analyses

Descriptive statistics were used to summarize sociodemographic and related characteristics: frequencies and percentages for categorical variables and means and standard deviations for continuous variables. We used population-averaged Poisson models (generalized estimating equations or GEE) [46] to estimate dichotomous outcomes (i.e., depression, anxiety), and zero-truncated Poisson models [47] to estimate count outcomes (i.e., Covid-19 knowledge, Covid-19 protective behaviors). Incidence rate ratios (IRR) were calculated based on pair-wise comparisons to estimate between group differences on primary outcomes at baseline, postintervention, and 2-month follow-up. Clustering of observations at the participant level were taken into account by specifying the clustering variable (participant ID) in the GEE models and by using robust standard errors in the truncated Poisson models [48]. We conducted an intention-to-treat analysis, in that all participants were included irrespective of their session attendance or completion of questionnaires. A complete-case analysis is generally not recommended due to loss of sample size and potential bias in the results [49]. Models were adjusted for demographic factors such as age, employment status, sexual/gender identity subgroup, education, and race/ethnicity. Two-sided p-values < 0.05 were considered statistically significant. All analyses were performed using Stata/SE 16.1 (Stata Corporation, College Station, TX).

## Results

### Enrollment and Intervention Exposure

Among individuals initially screened online (n = 229), 27 (11.8%) were ineligible; of these, n = 18 did not meet residency requirements and n = 9 did not meet sexuality/gender criteria. Among enrolled participants (n = 202), the majority (54.5%; n = 110) completed 3 sessions, 5 (2.5%) completed 2 sessions, 8 (4.0%) 1 session, and 79 (39.1%) no sessions. Fifty-four percent (n = 109) completed the postintervention survey, and 48% (n = 96) completed the follow-up survey (see Fig 1).

**Fig 1.**
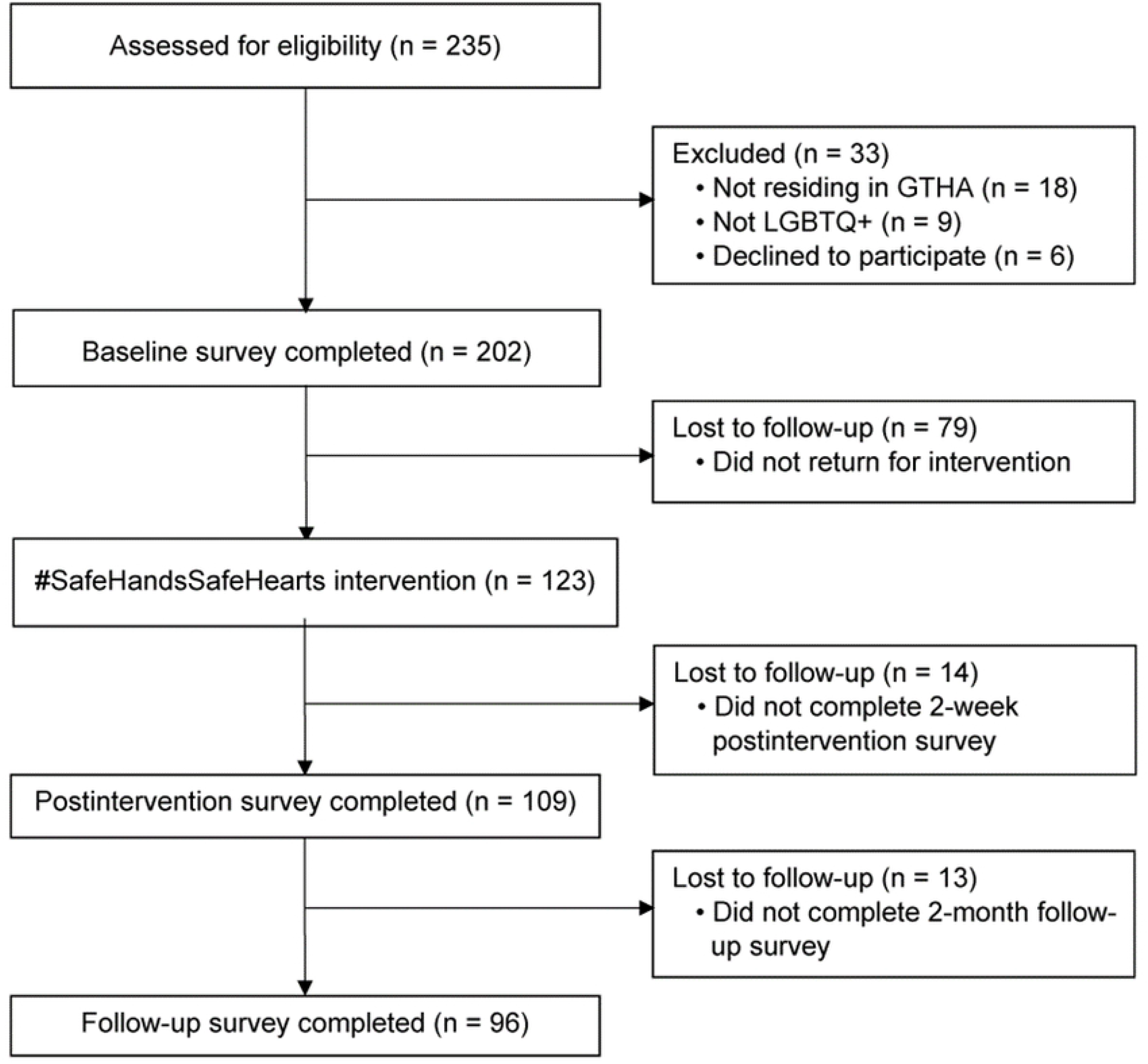
CONSORT diagram of participant flow in one-group quasi-experimental pretest-posttest design (#SafeHandsSafeHearts) Figure legend: GTHA, Greater Toronto and Hamilton Area; LGBTQ+, lesbian, gay, bisexual, transgender, queer, and other persons outside of heteronormative and cisgender identities

### Participant Characteristics

In total, 202 participants (median age, 27 years [IQR: 23-32]) were enrolled in the #SafeHandsSafeHearts trial. The majority (54%, n = 110) identified as cisgender LBWSW, nearly one-fifth (19.3%, n = 39) TNB, and one-quarter (26.2%, n = 53) cisgender GBMSM. Nearly one-third of participants (30.7%, n = 62) identified as African/Caribbean/Black, 30.2% (n = 61) South/East/Southeast Asian, 24.3% (n = 49) White, 8.9% (n = 18) Latinx/Hispanic, and 5.9% (n = 12) multiracial. Over two-thirds (67.3%, n = 136) had college-degree education and one-half (50.0%, n = 101) were unemployed. Table 1 shows participant demographic characteristics.

**Table 1.** Baseline demographic characteristics (N = 202)

| **Characteristic** | **n (%)** |
| --- | --- |
| **Race/ethnicity** |  |
| African/Caribbean/Black | 62 (30.7) |
| South/East/Southeast Asian | 61 (30.2) |
| Latinx/Hispanic | 18 (8.9) |
| White | 49 (24.3) |
| Multiracial | 12 (5.9) |
| **Gender** |  |
| Cisgender lesbian, bisexual, WSW | 110 (54.5) |
| Cisgender gay, bisexual, MSM | 53 (26.2) |
| Transgender or non-binary | 39 (19.3) |
| **Education** |  |
| None/Elementary school | 6 (3.0) |
| High school | 60 (29.7) |
| ≥ College/University | 136 (67.3) |
| **Employment** |  |
| Full-time | 47 (23.3) |
| Part-time | 54 (26.7) |
| Unemployed | 101 (50.0) |
| **Health Insurance** |  |
| Insured | 164 (81.2) |
| Uninsured | 38 (18.8) |

MSM, men who have sex with men; WSW, women who have sex with women

### Psychological Distress, Covid-19 Knowledge and Protective Behaviors

The baseline prevalence of depressive symptoms was 53.6% (95% CI 48.0%–59.3%), and of anxiety symptoms was 63.5% (95% CI 57.5%–69.5%). The mean score on Covid-19 knowledge at baseline was 6.73 (95% CI 6.66–6.80) and on protective behaviors was 14.75 (95% CI 14.27–15.23) (see Table 2).

**Table 2.** Effect of the intervention on primary outcomes: Predicted probabilities (%) or scores, and pairwise comparisons of change in % or score at 3 timepoints.

| Outcomes | Scores or %'s at 3 timepoints<br>(95% CI) |  |  | Pairwise comparisons<br>IRR (95% CI) |  |  |
| --- | --- | --- | --- | --- | --- | --- |
|  | *Baseline*<br>*(T0)* | *Post-intervention*<br>*(T1)* | *Follow-up*<br>*(T2)* | **T0 to T1** | **T0 to T2** | **T1 to T2** |
| **Depressive symptoms (%)** | 53.6%<br>(48.0, 59.3) | 40.0%<br>(32.4, 47.7) | 49.1%<br>(39.2, 58.9) | 0.746**<sup>a</sup><br>(0.612 - 0.911) | 0.915<br>(0.737 - 1.135) | 1.225<br>(0.951 - 1.579) |
| **Anxiety symptoms (%)** | 63.5%<br>(57.5, 69.5) | 52.9%<br>(44.5, 61.3) | 48.4%<br>(38.8, 58.0) | 0.834*<br>(0.710 - 0.979) | 0.763*<br>(0.615 - 0.945) | 0.915<br>(0.732 - 1.144) |
| **Covid-19 knowledge score (2–7)** | 6.73<br>(6.66, 6.80) | 6.68<br>(6.57, 6.80) | 6.67<br>(6.52, 6.82) | 0.992<br>(0.973 - 1.013) | 0.991<br>(0.967 - 1.016) | 0.998<br>(0.970 - 1.027) |
| **Covid-19 protective behaviors score (1–18)** | 14.75<br>(14.27, 15.23) | 15.48<br>(15.02, 15.94) | 16.02<br>(15.55, 16.49) | 1.049*<sup>b</sup><br>(1.002 - 1.099) | 1.086***<br>(1.036 - 1.138) | 1.035<br>(0.994 - 1.078) |

\*p<.05, \*\*p<.01, \*\*\*p<.001,
<sup>a</sup> IRRs can be interpreted as relative differences; an IRR of 0.746 signifies a 25.4% decrease in the prevalence of depression from baseline (T0) to postintervention (T1) (i.e., $0.746 - 1 = -.254$ or -25.4%)
<sup>b</sup> This signifies a 4.9% increase in the mean score of Covid-19 protective behaviors from baseline to postintervention ( $1.049 - 1 = .049$ or 4.9%) CI, confidence interval; IRR, incidence rate ratio

### Effect of the Intervention on Depression

The intervention led to statistically significant reductions in the prevalence of depressive symptoms by one-fourth (25.4%) from baseline to postintervention (IRR = 0.746; 95% CI 0.612–0.911, p<.01) (Table 2). Those who were employed reported significantly lower risk (IRR = 0.74; 95% CI .60–.90, p<.05) of depressive symptoms (Table 3). Higher scores on Covid-19 stress (IRR = 1.25; 95% CI, 1.13–1.39, p<.001) and loneliness (IRR = 1.24; 95% CI 1.16–1.32, p<.001) were associated with significantly greater risks of depressive symptoms (Table 2). Higher scores on resilience to traumatic stress were associated with a small but significantly lower risk of depressive symptoms (IRR = 0.97; 95% CI .94–.99, p<.001). Overall reduction in the prevalence of depression from baseline to 2-month follow-up was not statistically significant, nor was the difference between postintervention and follow-up prevalence of depressive symptoms statistically significant, suggesting that the effect of the intervention on depression was not retained.

**Table 3.** Predictors of depressive and anxiety symptoms among LGBTQ+ individuals in Toronto (n = 202)

| Variables | Depression |  |  | Anxiety |  |  |
| --- | --- | --- | --- | --- | --- | --- |
|  | IRR | SE | 95% CI | IRR | SE | 95% CI |
| **Time** (Ref. Baseline) |  |  |  |  |  |  |
| Postintervention | 0.75** | 0.08 | 0.61 - 0.91 | 0.83* | 0.07 | 0.71 - 0.98 |
| Follow-up | 0.91 | 0.10 | 0.74 - 1.13 | 0.76* | 0.08 | 0.62 - 0.94 |
| **Age** | 0.99 | 0.00 | 0.98 - 1.00 | 1.00 | 0.00 | 0.99 - 1.01 |
| **Employed** (Yes) | 0.74** | 0.07 | 0.61 - 0.90 | 0.84* | 0.07 | 0.70 - 0.99 |
| **Identity** (Ref. GBMSM) |  |  |  |  |  |  |
| LBWSW | 1.23 | 0.17 | 0.93 - 1.62 | 1.29* | 0.16 | 1.01 - 1.66 |
| TNB | 1.14 | 0.18 | 0.84 - 1.55 | 1.21 | 0.18 | 0.91 - 1.62 |
| **Education:** College (Ref. Up to higher secondary) | 1.00 | 0.10 | 0.83 - 1.21 | 1.04 | 0.10 | 0.86 - 1.25 |
| **Covid-19 stress score** | 1.25*** | 0.07 | 1.13 - 1.39 | 1.29*** | 0.07 | 1.16 - 1.43 |
| **Loneliness score** | 1.24*** | 0.04 | 1.16 - 1.32 | 1.14*** | 0.03 | 1.08 - 1.21 |
| **Resilience to traumatic experience score** | 0.97** | 0.01 | 0.94 - 0.99 | 1.00 | 0.01 | 0.98 - 1.02 |
| **Race** (Ref. White) |  |  |  |  |  |  |
| African/Caribbean/Black | 1.20 | 0.15 | 0.94 - 1.52 | 0.90 | 0.10 | 0.72 - 1.13 |
| South/East/SE Asian | 0.89 | 0.12 | 0.69 - 1.16 | 0.91 | 0.10 | 0.72 - 1.14 |
| Latinx/Hispanic | 1.11 | 0.27 | 0.69 - 1.79 | 1.18 | 0.21 | 0.84 - 1.66 |
| Multiracial | 1.16 | 0.16 | 0.89 - 1.51 | 1.17 | 0.19 | 0.85 - 1.61 |
| **Constant** | 0.15*** | 0.06 | 0.07 - 0.33 | 0.13*** | 0.05 | 0.07 - 0.26 |

\*p<.05, \*\*p<.01, \*\*\*p<.001
SE, standard error
GBMSM, gay, bisexual and men who have sex with men; LBWSW, lesbian, bisexual women who have sex with women; SE Asian, Southeast Asian; TNB, transgender or non-binary individuals

### Effect of the Intervention on Anxiety

The intervention led to statistically significant reductions in anxiety symptoms, with the prevalence reduced by 16.6% from baseline to postintervention (IRR = 0.834; 95% CI 0.710–0.970, p<.05) (Table 2). The prevalence of anxiety symptoms at 2-month follow-up was also significantly different from baseline (23.7% reduction: IRR = 0.763; 95% CI 0.615– 0.945, p<.05), with no significant difference from postintervention to follow-up (IRR = 0.915; 95% CI 0.732–1.144, p = .65); this suggests that the effect of the intervention on reducing anxiety was retained over time (Table 2). Those who were employed had a significantly lower risk (IRR = 0.84; 95% CI, .70–.99, p<.05) of anxiety symptoms (Table 3). Higher scores on Covid-19 stress (IRR = 1.29; 95% CI 1.16–1.43, p<.001) and higher scores on loneliness (IRR = 1.14; 95% CI 1.08–1.21, p<.001) were each associated with significantly greater risk of anxiety symptoms.

### Effect of the Intervention on Covid-19 Knowledge

There was no statistically significant increase in Covid-19 knowledge scores over time (Table 2). Higher scores on vaccine conspiracy beliefs (IRR = 0.994; 95% CI, 0.990– 0.998, p<.05) were inversely associated with Covid-19 knowledge scores over time (Table 4). Higher Covid-19 stress scores were positively associated with Covid-19 knowledge (IRR = 1.02; 95% CI, 1.005–1.04, p<.05) over time.

**Table 4.** Predictors of Covid-19 knowledge and protective behavior scores in Toronto (n = 202)

| Variables | Covid-19 Knowledge Score |  |  | Covid-19 Behavior Score |  |  |
| --- | --- | --- | --- | --- | --- | --- |
|  | IRR | SE | 95% CI | IRR | SE | 95% CI |
| **Time (Ref. Baseline)** |  |  |  |  |  |  |
| Postintervention | 0.99 | 0.01 | 0.97 - 1.01 | 1.05* | 0.02 | 1.00 - 1.10 |
| Follow-up | 0.99 | 0.01 | 0.97 - 1.02 | 1.09*** | 0.03 | 1.04 - 1.14 |
| **Age** | 1.00 | 0.00 | 1.00 - 1.00 | 1.00 | 0.00 | 1.00 - 1.00 |
| **Employed (Yes)** | 0.99 | 0.01 | 0.97 - 1.01 | 1.03 | 0.02 | 0.99 - 1.07 |
| **Identity (Ref. GBMSM)** |  |  |  |  |  |  |
| LBWSW | 1.00 | 0.01 | 0.98 - 1.02 | 1.03 | 0.03 | 0.98 - 1.09 |
| TNB | 1.01 | 0.01 | 0.98 - 1.03 | 1.02 | 0.04 | 0.95 - 1.10 |
| **Education: College (Ref. up to higher secondary)** | 1.02* | 0.01 | 1.00 - 1.04 | 0.97 | 0.02 | 0.93 - 1.02 |
| **Vaccine conspiracy beliefs score** | 0.99** | 0.00 | 0.990 - 0.998 | 1.00 | 0.00 | 0.99 - 1.00 |
| **Covid-19 stress score** | 1.02* | 0.01 | 1.005 - 1.04 | 1.03* | 0.01 | 1.00 - 1.05 |
| **Covid-19 knowledge score** |  |  |  | 1.06*** | 0.02 | 1.03 - 1.10 |
| **Vaccinated with ≥1 dose (Yes)** |  |  |  | 0.93* | 0.03 | 0.89 - 0.99 |
| **Race (Ref. White)** |  |  |  |  |  |  |
| African/Caribbean/Black | 1.02 | 0.01 | 1.00 - 1.05 | 1.06* | 0.03 | 1.00 - 1.12 |
| South/East/SE Asian | 1.02* | 0.01 | 1.00 - 1.05 | 1.05* | 0.03 | 1.00 - 1.11 |
| Latinx/Hispanic | 1.04** | 0.02 | 1.01 - 1.07 | 1.02 | 0.05 | 0.93 - 1.13 |
| Multiracial | 0.99 | 0.03 | 0.95 - 1.04 | 1.08* | 0.04 | 1.02 - 1.15 |
| **Vaccinated with ≥1 dose (Yes)** | 1.01 | 0.01 | 0.98 - 1.03 |  |  |  |
| **Constant** | 6.58*** | 0.17 | 6.25 - 6.92 | 9.01*** | 1.27 | 6.84 - 11.87 |

\*p<.05, \*\*p<.01, \*\*\*p<.001

### Effect of the Intervention on Covid-19 Protective Behaviors

The intervention led to a statistically significant increase in Covid-19 protective behavior scores over time, from baseline to postintervention (4.9% increase: IRR = 1.049; 95% CI 1.002–1.099, p<.05), and from baseline to 2-month follow-up (8.6% increase: IRR = 1.086; 95% CI 1.036–1.138, p<.001) (Table 2). There was no significant change in Covid-19 protective behavior scores from postintervention to 2-month follow-up (IRR = 1.035; 95% CI 0.994–1.078, p=.06), suggesting that the effect of the intervention was retained. Those who reported Covid-19 vaccination (one or two doses) showed small but statistically significantly differences in lower scores on Covid-19 protective behaviors (IRR = 0.93; 95% CI 0.89–0.99, p<.05) over time (Table 4).

### Acceptability of the Intervention

Evaluation forms were submitted online by 72% of those who engaged in intervention counselling sessions. Overall, participants indicated feeling “very satisfied” in 84% (141/168) of sessions evaluated and “very satisfied” with the duration of the session in 79% (133/168) of sessions evaluated. In 86% (145/168) of sessions evaluated, participants indicated they were “very helpful” in “improving your emotional wellness or mental health.”

## Discussion

This study provides evidence of the effectiveness of a novel community-based, peer-delivered eHealth intervention (#SafeHandsSafeHearts) in reducing psychological distress and increasing Covid-19 protective behaviors among ethnoracially diverse LGBTQ+ individuals. Despite the largescale failure of public health systems to systematically report sexual orientation and gender identity information in relation to Covid-19 [50], increasing evidence indicates the disproportionate impact of Covid-19 and pandemic-related public health responses on health and mental health outcomes among LGBTQ+ people in Canada [17], the US [51,52], and Western Europe [53]. To that end, this is the first intervention of which we are aware to demonstrate effectiveness in mitigating pandemic-related psychosocial and behavioral risks among LGBTQ+ people.

Importantly, even among research and interventions designed to support broader LGBTQ+ health, there is substantially lesser focus on individuals who occupy intersectional marginalized identities on the basis of sexual and gender minority and ethnoracial minority status, such as LGBTQ+ Black, Latinx, other people of color, and women [12,14,50]. Our successful enrollment of a sample of LGBTQ+ people, a majority of whom were Black and other people of color, is both notable and apropos of the increased risk of Covid-19 infection and worser outcomes among these populations [8]. The successful implementation of #SafeHandsSafeHearts during a global public health emergency, with extensive rolling lockdowns and stay-at-home orders in the GTHA [21], further supports the feasibility and ecological validity of the intervention.

As hypothesized, the intervention led to a significant reduction in the prevalence of symptoms indicative of clinically significant depression and anxiety from baseline to postintervention. The high baseline prevalence of depressive and anxiety symptoms itself exemplifies the extreme pandemic-related distress among ethnoracially diverse LGBTQ+ individuals, many of whom were enrolled before Covid-19 vaccination was available to them, beginning in June 2021 [54].

The significant effects of the intervention in reducing the prevalence of depression and anxiety support the success of the MI-based and psychoeducational approach, as well as the eHealth modality. The decidedly nonjudgmental stance of MI may be especially appropriate in working with sexual and gender minority and ethnic minority communities, more so amid a pandemic. These communities may experience ambivalence or alienation in the context of historically justified medical mistrust [55,56], moreover, in response to public health interventions that were not designed with community input nor with their communities in mind [57]. The use of eHealth, as well as telehealth and other online modalities with low barriers to access, also may be particularly important for LGBTQ+, ethnoracial minority, and other marginalized populations during a pandemic; closures of familiar community-based services may reduce rather than increase access to culturally competent health and mental health services despite their heightened vulnerability [53].

Despite the postintervention reductions in depression and anxiety, the increase in depressive symptomology from post-intervention to 2-month follow up suggests that future research using an RCT design should assess the intervention using biweekly versus weekly sessions, more than 3 sessions, and/or the use of eHealth booster sessions [58] to determine if any of these may support retention of reductions in depression over time. The retention of intervention effects in reducing anxiety at 2-months post-intervention during an ongoing pandemic suggests the potential for more sustained impact on depression.

We also identified statistically significant associations between secondary measures of loneliness and Covid-19 stress, respectively, and depression and anxiety outcomes. The intervention may have helped to abate these risk factors, potentially reflecting benefits of a peer-based approach in providing psychosocial support for marginalized communities in the context of systemic stigma and discrimination [59]. LGBTQ+ individuals may be particularly vulnerable to psychological distress during lockdowns as a result of different family configurations than heterosexual individuals on whom public health responses are normed, and the closure of community spaces that provide LGBTQ+-affirmative and culturally competent support [60,61].

Notably, employment exerted significant protective effects against both depression and anxiety; this corroborates pathways through which economic marginalization contributes to vulnerability in a pandemic [18,50]. The extensive job loss reported among our sample is substantiated by Toronto government data indicating 50% higher rates of pandemic-related unemployment among Black and Asian versus other ethnoracial groups [62], as well as studies demonstrating vulnerability and stress associated with pandemic-related job loss among LGBTQ+ individuals in Canada [18] and the U.S. [20,50]. Structural interventions to promote job security and retention among ethnoracial and sexual and gender minorities, communities disproportionately represented in service industries [3], including provision of paid sick leave [63] and broader employment antidiscrimination measures, may exert substantial salubrious effects on mental health in a public health emergency.

Our findings also identify relatively high adherence to public health-recommended protective behaviors at baseline, including physical distancing and mask-wearing, as well as handwashing. The intervention had a small but sustained positive effect on protective behaviors, with increases retained at 2-month follow-up. The uniformly high Covid-19 knowledge demonstrated at baseline may be attributable to daily public health messaging about Covid-19 delivered through multiple channels in the Toronto area; this may have impeded our ability to detect improvements in Covid-19 knowledge over time.

## Strengths and Limitations

Study results should be understood in the context of limitations. First, the quasi-experimental pretest-posttest design precludes the ability to determine causal associations; however, the temporality of the intervention, and use of pretest, posttest, and follow-up surveys, support the impact of the intervention on the outcomes assessed and account for possible decay in intervention effects over time [64]. Second, self-reporting of depression and anxiety symptoms, and use of a brief assessment instrument, might result in bias; however, we implemented widely used screening tools with established validity and reliability [35].

Third, while over 60% of individuals who screened into the study completed 1 or more—89% of these, all 3—counseling sessions, session nonattendance may reflect the challenges of participation and retention during a pandemic. We anticipated constraints to engaging in online counseling owing to lack of privacy, and not having a personal mobile device or broadband internet access. To that end, we made arrangements with local CBOs to provide free PC/internet access on site; however, ongoing lockdowns and concerns about infection may have precluded their utilization. Nevertheless, even with the inclusion of participants who did not attend one or more sessions in our analyses, we identified intervention effectiveness on three of four primary outcomes. Fourth, lack of retention of significant reductions in depression, similarly identified in other brief MI-based interventions [32], suggests the need to test #SafeHandsSafeHearts with additional eHealth sessions over a longer duration. Finally, study results may not be generalizable to all ethnoracially diverse LGBTQ+ people in Toronto or elsewhere. However, we designed eligibility criteria to include a broad range of self-identifications among sexual and gender minorities (i.e., not only self-identified gay or lesbian or trans individuals) and we were successful in recruitment of a racially and ethnically diverse LGBTQ+ sample during a pandemic.

## Conclusion

We demonstrated the preliminary effectiveness of an innovative, brief eHealth intervention in reducing psychological distress and increasing protective behaviors among ethnoracially diverse LGBTQ+ individuals during the Covid-19 pandemic. The successful implementation of #SafeHandsSafeHearts through partnership and strengthening of community-based health services, and by trained peer counselors with ongoing clinical supervision, most of whom mirrored the sample demographics, supports feasibility and the need for further evaluation of the #SafeHandsSafeHearts model. Overall, this study also affirms the critical importance of meaningful engagement of ethnoracially diverse LGBTQ+ communities in pandemic preparedness, and public health intervention design and implementation.

## Data Availability

The data will be held in a public repository and will only be available after acceptance.

## Authorship Confirmation/Contribution Statement

P.A.N.: conceptualization, methodology, formal analysis, writing—original draft, writing—review and editing, funding acquisition. V.C.: conceptualization, methodology, formal analysis, writing—review and editing. N.M.: methodology, supervision. C.C.W.: conceptualization, methodology. W.T.: resources, supervision. S.T.: project administration. S.R.: software, data curation. J.F.: supervision. S.S.: project administration. P.A.: data curation. M.A.: supervision, project administration. All authors reviewed and approved the final article before submission.

## Acknowledgements

The authors would like to acknowledge our lead community partner, Women’s Health in Women’s Hands Community Health Centre, for their collaboration in implementing the study. We also thank our dedicated online peer counsellors and study staff: Chantel Aboagye-Mensah, Celeste Bilbao, Rachel Cheung, Denise Frans, Manvinder Gill, Rae Paul, Ali Pearson, Monte-Angel Richardson, Jora Shacter, Jena Taylor-Hunter, Monica Vu, Alisha Williams, and Alexandra Wright.

